# Association between Renal Function and Retinal Neurodegeneration in Chinese Patients with Type 2 Diabetes Mellitus

**DOI:** 10.1101/2020.11.22.20236331

**Authors:** Xia Gong, Wei Wang, Wangting Li, Ling Jin, Lanhua Wang, Jie Meng, Kun Xiong, Yuting Li, Xiao Han, Xiaoling Liang, Yizhi Liu, Wenyong Huang

## Abstract

**Background:** Chronic kidney disease (CKD) and diabetic retinopathy (DR) are two serious complications of diabetes. However, the association between retinal neurodegeneration in DR and renal function decline is still unclear. Our objective was to evaluate the association by measure estimated glomerular filtration rate (eGFR), macular ganglion cell–inner plexiform layer (GC–IPL) and ganglion cell complex (GCC) thickness in patients with type 2 diabetes mellitus (T2DM).

**Methods:** We analyzed the baseline data of the Guangzhou Diabetic Eye Study. T2DM patients from communities in Guangzhou were enrolled and all participants went through ophthalmic and general examinations. The thickness of the macular GC–IPL and GCC in their right eyes were measured by swept-source optical coherence tomography. CKD was defined as eGFR < 60 mL/min/1.73 m^2^.

**Results:** 1,309 patients were included (mean age 64.4 ± 7.6 years, 59.1% female), and fifty-eight (4.4%) of them had CKD. Average macular GC-IPL thickness was significantly thinner in CKD patients (96.5 ± 9.1 μm) than non-CKD patients (101.3 ± 9.2 μm) (*p* < 0.01). Average macular GCC thickness was also significantly thinner in CKD patients (123.5 ± 13.2 μm) than non-CKD patients (129.9 ± 12.8 μm) (*p* < 0.01). The significant thinning of macular GC-IPL and GCC thickness presented in every gird in macula (all, p < 0.05) except for central grid (p ≥ 0.05). In the patients without DR, the eGFR was linearly correlated with the average macular GC–IPL thickness (β = 0.07 [95% CI 0.02–0.12], *p* < 0.01) and GCC thickness (β = 0.09 [95% CI 0.03–0.16], *p* < 0.01) after adjustment for age, sex, axial length, intraocular pressure and combination of hypertension. However, no linear correlation was found between eGFR and macular GC-IPL or GCC thickness in DR patients.

**Conclusions:** Renal function decreases is associated with the thinning of the macular GC–IPL and GCC in T2DM patients, suggesting the potential value of ganglion cell lose to detect early function decline in the kidney in diabetic patients, especially in patients without DR.

## 1. Background

Chronic kidney disease (CKD) is one of the major complications in diabetic patients. CKD is not only a cause of renal failure, but also an independent risk factor for cardiovascular disease, cognitive dysfunction, and all-cause mortality. With the increasing diabetic patients, the number of patients with CKD has increased dramatically in recent decades and has become one of the greatest global health challenges of our time.^1,2^ However, most CKD patients were diagnosed after renal function obvious decline. Thus it is important to detect renal impairment in the early stage of CKD development.^2^

Diabetic retinopathy (DR) is another major complications in diabetic patients.^3^ In many countries, DR, which develops in approximately 20–40% of diabetic patients,^4^ is the most common cause of avoidable blindness in the working-age population.^5^ Diabetic patients are suggested to take regular screening for DR.

Because both CKD and DR are microvascular complications of diabetes, researchers suggested that CKD and DR may be closely linked.^6^ The earliest clinical feature of DR is vascular changes, while retinal ganglion cell–inner plexiform layer (GC–IPL) thickness and ganglion cell complex (GCC) thickness are known to be early indicators of hyperglycemia-induced neurodegeneration prior to the detectability of clinical microvascular abnormalities.^7-9^ Hyperglycemia-induced neurodegeneration affects primarily the retinal ganglion cell nuclei and dendrites. This then affects the axons of the ganglion cell, and this is manifested as the diffuse thinning of the retinal GC–IPL followed by the thinning of the retinal nerve fiber layer (RNFL).^7^ Since GCC is the combination of GC-IPL and RNFL, it will become thinner in this process. Moreover, researchers suggested that neurdegeneration in retina may associated with renal function decline in diabetic patients.^10^

However, the association of the important parameters of neurodegeneration in DR development, including the thickness of the macular GC–IPL and GCC, with the eGFR (the most valuable indicator of overall renal function),^11^ has not been elucidated. To fill this gap, we analyzed the baseline data of the Guangzhou Diabetic Eye Study to evaluate the associations. This study is the first to use swept-source optical coherence tomography (SS-OCT) to investigate the association between retinal ganglion cell loss and renal function in T2DM patients in China.

We present the following article/case in accordance with the Strengthening the Reporting of Observational Studies in Epidemiology (STROBE) reporting checklist.

## 2. Methods

### 2.1. Subjects

The Guangzhou Diabetic Eye Study is a prospective study conducted in the Zhongshan Ophthalmic Center, which is affiliated with Sun Yat-Sen University in Guangzhou, China. The study protocol was approved by the Zhongshan Ophthalmic Center Ethics Committee. Written consent was obtained from all participants before they entered the study. This article analyzed its baseline data collected from October 2018 to April 2019

The study recruited 1,473 T2DM patients between the ages of 30 and 80. They were already registered in the health care systems in the communities near the Zhongshan Ophthalmic Center. Patients with any evidence of the following conditions were excluded: (1) best corrected visual acuity (BCVA) worse than 20/200, axial length > 30 mm or unmeasurable, spherical equivalent (SphE) ≤ –12.0 degrees, astigmatism > 4 degrees, or intraocular pressure > 21 mmHg in the study eye; (2) except DR, other combined eye diseases that could affect retinal thickness in the study eye, such as glaucoma, age-related macular degeneration, and retinal detachment; (3) surgery or invasive treatment or laser treatment history on the study eye; (4) severe systemic diseases, such as uncontrolled hypertension, severe cardiovascular and cerebrovascular disease, malignant tumors, and nephritis; (5) general surgery history, such as heart bypass, thrombolysis, and kidney transplantation; (6) cognitive disorders or mental illness that would hinder the patient’s cooperation with tests; and (7) inability to obtain clear fundus or SS-OCT images because of refractive media opacity or noncooperation.

We excluded 40, 47, 77 patients for unqualified general condition, eye condition or image quality, respectively.

### 2.2. General Information and Laboratory Measurements

Upon enrollment in the study, general information, including age, sex, diabetes duration, other combined systemic chronic diseases, medication compliance, and smoking and drinking histories, were collected via questionnaires.

Height and weight were manually measured. Body mass index (BMI) was defined as weight (kg) divided by the square of height (m), and a patient with a BMI of 23–27.5 kg/m^2^ or >27.5 kg/m^2^ was considered to be overweight or obese, respectively.^12^

Each patient’s systolic blood pressure (SBP) and diastolic blood pressure (DBP) were measured by electronic sphygmomanometer (Hem-907, Omron, Kyoto, Japan). The mean artery pressure (MAP) was calculated to be the sum of 1/3 SBP and 2/3 DBP.

Blood and urine samples were collected by experienced nurses. Serum creatinine, hemoglobin A1c (HbA1c), total cholesterol (TC), high-density lipoprotein cholesterol (HDL-C), low-density lipoprotein cholesterol (LDL-C), triglycerides (TG), C-reactive protein, and microalbuminuria were measured. The eGFR was calculated with the Xiangya formula, which has been reported to be the most accurate eGFR formula for the Chinese population.^13^ CKD was defined as an eGFR lower than 60/min/1.73 m^2^.^14^

Participants who had abnormal observations or intake of related drugs were defined as hypertension or hyperlipidemia.

### 2.3. Eye Examination

Each participant underwent a comprehensive ocular examination that included the best correct visual acuity (BCVA) test (Early Treatment Diabetic Retinopathy Study [ETDRS] LogMAR visual acuity chart, Precision Vision, Artesia, USA), refraction powers examination (auto refractometer, KR-8800, Topcon, Tokyo, Japan), intraocular pressure (IOP) measurement (non-contact tonometer, CT-1, Topcon, Tokyo, Japan), slit lamp examination, and ocular biometric measurements (Lenstar LS900, Haag-Streit, Koeniz, Switzerland). The SphE was calculated as the sphere degree plus 1/2 cylinder degree. The standard ETDRS 7-field fundus photography (CR-2, Canon, Tokyo, Japan) was obtained after the pupil was dilated with 3 drops of tropicamide phenylephrine (Mydorin, Santen, Osaka, Japan). The images were graded by two well-trained graders in accordance with the English National Health Service Diabetic Eye Screening Programme Grading Classification Standard.^15^

### 2.4. Swept-Source Optical Coherence Tomography

A SS-OCT device (DRI OCT Triton, Topcon, Tokyo, Japan) was used to scan the fovea area. A three-dimensional (3D) imaging scan protocol of native software (10.15.003.01) was used to evaluate a 6 × 6 mm region of the macula. The scans were centered automatically and confirmed by a fundus camera that was integrated into the instrument. In accordance with the ETDRS standard, three circles with diameters of 1, 3, 6mm divided the fovea into a central grid, an inner ring and an outer ring; then inner ring and outer ring were divided into four quadrants, including superior, inferior, nasal and temporal quadrants. The thickness profiles of the GC–IPL and GCC layers were automatically produced, and thickness of GC-IPL or GCC in each grid was automatically reported.

The examinations were performed by an experienced examiner with no knowledge of the study protocol. Each participant’s measurements were obtained through dilated pupil. The exclusion of unqualified images, including out-of-focus images or those with motion artifacts or segmentation algorithm failures, was performed at a different time from that of image acquisition.

### 2.5. Statistical Analysis

The data from each participant’s right eye was analyzed. The difference in characteristics between the CKD patients and non-CKD patients was evaluated through the independent samples *t*-test for the normally distributed data, Spearman’s rank-sum test for the non-normally distributed data, and chi-square test for the qualitative data respectively. Univariate and multivariate linear regressions were used to evaluate the relationships between the eGFR and macular GC–IPL and GCC thickness. Model 1 of the multivariable linear regression was adjusted for the factors known to be relevant to macular GC–IPL and GCC thickness. These factors included age, sex, and axial length.^16^ Model 2 was further adjusted for the parameters that were associated with macular GC–IPL and GCC thickness in the univariable linear regression. These factors included hypertension and IOP. A *p*-value of <0.05 was considered significant.

## 3. Results

A total of 1,309 diabetic patients (59.1% female) were included in the final analysis. The demographic and clinical characteristics are presented in Table 1. They had an average age of 64.4 ± 7.6 years (mean ± standard deviation [*SD*]). The median (interquartile range) diabetic duration was 7.0 (3.0–13.0) years. The proportion of overweight (51.6%), obesity (16.8%), hypertension (56.5%), or hyperlipidemia (47.2%) were high. Participants had a vision of average LogMAR BCVA (SD) 0.2 (0.1). Among the participants, 1,094 (83.6%) had no signs of DR and 215 (16.4%) had DR.

**Table 1.** Participant’s Characteristics Stratified by the Presence of Chronic Kidney Diseases.

| Characteristics | All participants | CKD | Non-CKD | P-value |
| --- | --- | --- | --- | --- |
| No. (eyes) | 1309 | 58 | 1251 |  |
| Age, mean (SD), year | 64.4 (7.6) | 70.7 (6.6) | 64.1 (7.5) | **<0.001**<sup>†</sup> |
| Sex |  |  |  | 0.684<sup>§</sup> |
| Male, % | 40.9 | 37.9 | 41.0 |  |
| Female, % | 59.1 | 62.1 | 59.0 |  |
| Body mass index, mean (SD), kg/m<sup>2</sup> | 24.6 (3.2) | 25.6 (3.3) | 24.5 (3.2) | **0.018**<sup>†</sup> |
| Diabetes: |  |  |  |  |
| Duration of diabetes, median (IQR), year | 7.0 (3.0-13.0) | 11.0 (5.0-17.0) | 7.0 (3.0-13.0) | **0.002**<sup>‡</sup> |
| HbA1c, mean (SD), % | 7.0 (1.5) | 6.9 (1.5) | 7.0 (1.5) | 0.847<sup>†</sup> |
| Use of insulin treatment, % | 20.9 | 36.2 | 20.1 | **0.019**<sup>§</sup> |
| Diabetic retinopathy, % | 16.4 | 23.2 | 16.4 | 0.199<sup>§</sup> |
| Blood pressure: |  |  |  |  |
| Hypertension, % | 56.5 | 91.4 | 54.9 | **<0.001**<sup>§</sup> |
| SBP, mean (SD), mmHg | 134.6 (19.0) | 138.4 (19.2) | 134.4 (19.0) | 0.123<sup>†</sup> |
| DBP, mean (SD), mmHg | 70.0 (11.8) | 70.8 (9.3) | 69.6(11.9) | 0.460<sup>†</sup> |
| MAP, mean (SD), mmHg | 92.0 (11.9) | 93.3 (11.4) | 92.0 (11.9) | 0.405<sup>†</sup> |
| Blood lipid: |  |  |  |  |
| Hyperlipidemia, %: | 47.2 | 69.0 | 46.2 | **0.001**<sup>§</sup> |
| TG, mean (SD), mmol/L | 2.3 (1.6) | 2.6 (2.0) | 2.3 (1.6) | 0.121<sup>†</sup> |
| LDL-C, mean (SD), mmol/L | 3.1 (0.9) | 3.2 (1.1) | 3.1 (0.9) | 0.523<sup>†</sup> |
| HDL-C, mean (SD), mmol/L | 1.3 (0.4) | 1.2 (0.4) | 1.3 (0.4) | 0.107<sup>†</sup> |
| TC, mean (SD), mmol/L | 4.8 (1.0) | 5.0 (1.2) | 4.8 (1.0) | 0.266<sup>†</sup> |
| C-reactive protein, median (IQR), mg/dL | 1.4 (0.7- 2.7) | 2.0 (1.0- 3.8) | 1.4 (0.7- 2.6) | **0.027<sup>‡</sup>** |
| Renal function: |  |  |  |  |
| eGFR, mean (SD), mL/min/1.73 m<sup>2</sup> | 76.2 (10.7) | 54.2 (5.2) | 77.2 (9.8) | **<0.001<sup>†</sup>** |
| Serum creatinine, mean (SD), µmol/L | 71.4 (21.6) | 123.8 (35.5) | 69.0 (17.3) | **<0.001<sup>†</sup>** |
| Microalbuminuria, median (IQR), mg/L | 0.9 (0.3-2.8) | 5.4 (1.4-21.0) | 0.9 (0.3- 2.7) | **<0.001<sup>‡</sup>** |
| Eye examination: |  |  |  |  |
| LogMAR BCVA, mean (SD) | 0.2 (0.1) | 0.3 (0.1) | 0.2 (0.1) | **0.001<sup>†</sup>** |
| IOP, mean (SD), mmHg | 15.9 (2.5) | 15.0 (2.7) | 15.9 (2.4) | **0.011<sup>†</sup>** |
| Axial length, mean (SD), mm | 23.5 (1.1) | 23.5 (1.1) | 23.5 (1.1) | 0.817<sup>†</sup> |
| SphE, mean (SD), degree | 0.2 (2.2) | 0.2 (2.0) | 0.2(2.2) | 0.903<sup>†</sup> |

Abbreviations: CKD, chronic kidney diseases; SD, standard deviation; IQR, interquartile range; SBP, systolic blood pressure; DBP, diastolic pressure; MAP, mean artery pressure; TG, triglyceride; LDL-C, low density lipoprotein cholesterol; HDL-C, high density lipoprotein cholesterol; TC, total cholesterol; eGFR, estimated glomerular filtration rate; BCVA, best corrected visual acuity; IOP, intraocular pressure; SphE, spherical equivalent.
CKD defined as eGFR<60 mL/ min/1.73 m<sup>2</sup>.
<sup>†</sup> Independent two-sample *t* test
<sup>‡</sup> Spearman's rank-sum test
<sup>§</sup> Chi-square test

Fifty-eight (4.4%) participants had CKD. CKD patients were older than non-CKD patients (70.7 ± 6.6 years for CKD, 64.1 ± 7.5 years for non-CKD, *p* < 0.01). In addition, CKD patients had longer diabetes duration, higher BMI, higher C-reactive protein, higher microalbuminuria, worse BCVA, and, unexpectedly, lower IOP. The CKD patients were more likely to have hypertension, hyperlipidemia and have a requirement for insulin treatment (all *p* < 0.05).

Table 2 shows the macular GC–IPL and GCC thickness in CKD patients and non-CKD patients. The average macular GC–IPL was thinner in CKD patients than non-CKD patients, with measurements of 101.3 ± 9.2 μm for non-CKD, 96.5 ± 9.1 μm for CKD, respectively (*p* < 0.01). The average macular GCC was also thinner in CKD patient than non CKD patients, with measurements of 129.9 ± 12.8 μm for non-CKD, 123.5 ± 13.2 μm for CKD, respectively (*p* < 0.01). The macular GC–IPL and GCC in the CKD patients were significantly thinner than non-CKD patients in almost every grid (all, *p* < 0.05), except for central grid (*p* ≥ 0.05).

**Table 2.**
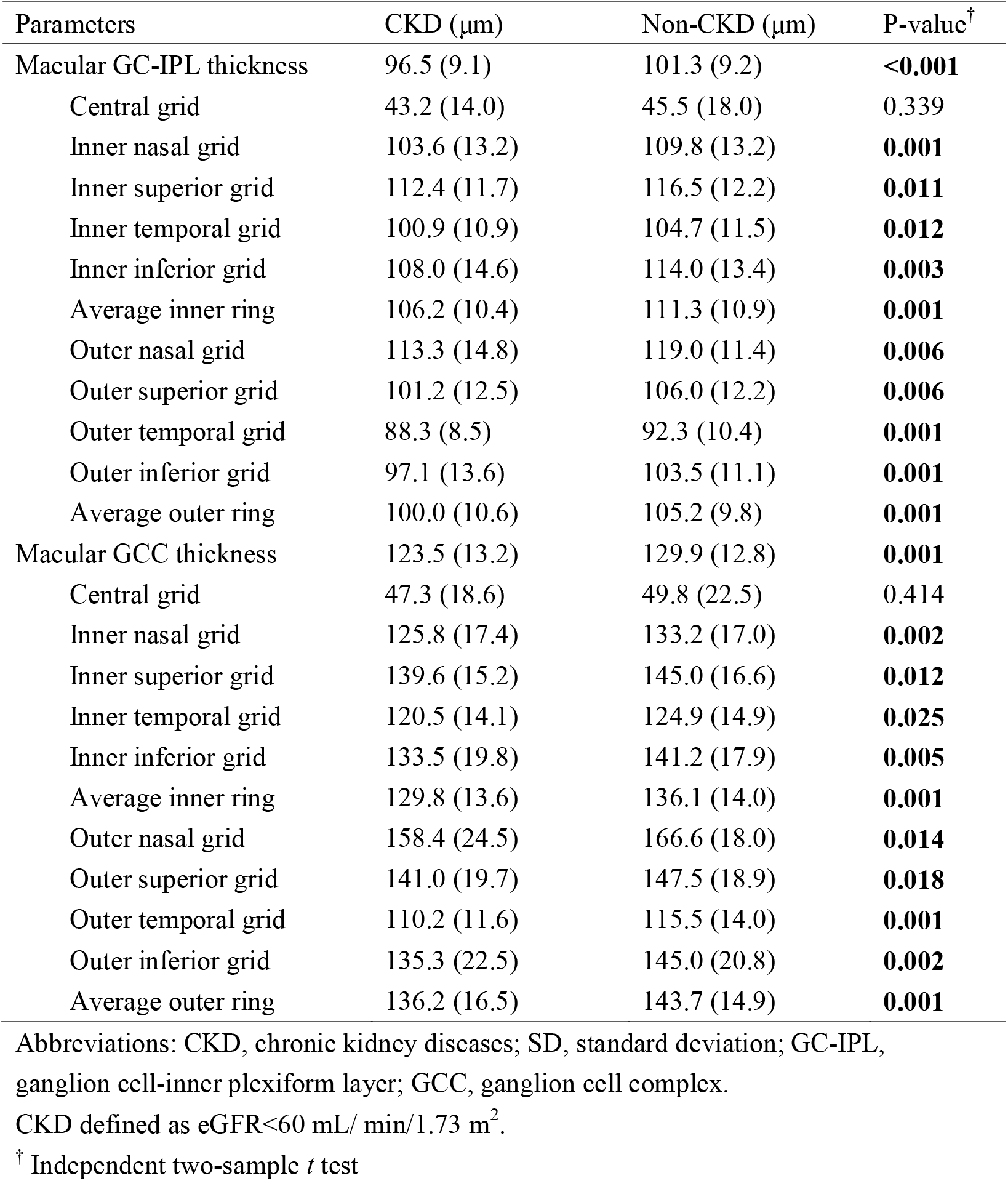
Thickness of Macular Ganglion Cell-inner Plexiform Layer and Ganglion Cell Complex Stratified by the Presence of Chronic Kidney Diseases.

| Parameters | CKD ($\mu\text{m}$) | Non-CKD ($\mu\text{m}$) | P-value<sup>†</sup> |
| --- | --- | --- | --- |
| Macular GC-IPL thickness | 96.5 (9.1) | 101.3 (9.2) | **<0.001** |
| Central grid | 43.2 (14.0) | 45.5 (18.0) | 0.339 |
| Inner nasal grid | 103.6 (13.2) | 109.8 (13.2) | **0.001** |
| Inner superior grid | 112.4 (11.7) | 116.5 (12.2) | **0.011** |
| Inner temporal grid | 100.9 (10.9) | 104.7 (11.5) | **0.012** |
| Inner inferior grid | 108.0 (14.6) | 114.0 (13.4) | **0.003** |
| Average inner ring | 106.2 (10.4) | 111.3 (10.9) | **0.001** |
| Outer nasal grid | 113.3 (14.8) | 119.0 (11.4) | **0.006** |
| Outer superior grid | 101.2 (12.5) | 106.0 (12.2) | **0.006** |
| Outer temporal grid | 88.3 (8.5) | 92.3 (10.4) | **0.001** |
| Outer inferior grid | 97.1 (13.6) | 103.5 (11.1) | **0.001** |
| Average outer ring | 100.0 (10.6) | 105.2 (9.8) | **0.001** |
| Macular GCC thickness | 123.5 (13.2) | 129.9 (12.8) | **0.001** |
| Central grid | 47.3 (18.6) | 49.8 (22.5) | 0.414 |
| Inner nasal grid | 125.8 (17.4) | 133.2 (17.0) | **0.002** |
| Inner superior grid | 139.6 (15.2) | 145.0 (16.6) | **0.012** |
| Inner temporal grid | 120.5 (14.1) | 124.9 (14.9) | **0.025** |
| Inner inferior grid | 133.5 (19.8) | 141.2 (17.9) | **0.005** |
| Average inner ring | 129.8 (13.6) | 136.1 (14.0) | **0.001** |
| Outer nasal grid | 158.4 (24.5) | 166.6 (18.0) | **0.014** |
| Outer superior grid | 141.0 (19.7) | 147.5 (18.9) | **0.018** |
| Outer temporal grid | 110.2 (11.6) | 115.5 (14.0) | **0.001** |
| Outer inferior grid | 135.3 (22.5) | 145.0 (20.8) | **0.002** |
| Average outer ring | 136.2 (16.5) | 143.7 (14.9) | **0.001** |

Abbreviations: CKD, chronic kidney diseases; SD, standard deviation; GC-IPL, ganglion cell-inner plexiform layer; GCC, ganglion cell complex.
CKD defined as eGFR<60 mL/ min/1.73 m<sup>2</sup>.
<sup>†</sup> Independent two-sample *t* test

Table 3 presents the results of the univariate linear regression analysis between participants’ characteristics and thickness of macular GC-IPL and GCC. Not only the eGFR but also age, sex, axial length, IOP, and combination of hypertension or DR were significantly linearly correlated with macular GC–IPL and GCC thickness (all, *p* < 0.05).

**Table 3.** Univariate Linear Regression Analysis of the Relationship between Characteristics and Thickness of Macular Ganglion Cell-inner Plexiform Layer or Ganglion Cell Complex.

| Characteristics | Macular GC-IPL thickness |  | Macular GCC thickness |  |
| --- | --- | --- | --- | --- |
|  | Coefficient (95%CI) | P-value | Coefficient (95%CI) | P-value |
| Age, year | -0.31 (-0.37, -0.24) | **<0.001** | -0.39 (-0.48, -0.30) | **<0.001** |
| Gender, male | 1.44 (0.42, 2.45) | **0.006** | 1.82 (0.39, 3.24) | **0.012** |
| Body mass index, kg/m<sup>2</sup> | -0.02 (-0.17, 0.14) | 0.802 | 0.01 (-0.21, 0.22) | 0.952 |
| Duration of diabetes, year | -0.03 (-0.10, 0.05) | 0.464 | -0.03 (-0.13, 0.08) | 0.626 |
| HbA1c, % | 0.21 (-0.13, 0.56) | 0.221 | 0.32 (-0.16, 0.80) | 0.193 |
| Use of insulin treatment | 0.56 (-0.67, 1.79) | 0.374 | 0.93 (-0.80, 2.65) | 0.292 |
| Diabetic retinopathy | 2.80 (1.46, 4.15) | **<0.001** | 4.09 (-2.20, 5.97) | **<0.001** |
| Hypertension | -1.80 (-2.80, -0.80) | **<0.001** | -2.15 (-3.57, -0.74) | **0.003** |
| SBP, mmHg | -0.02 (-0.05, -0.00) | 0.091 | -0.01 (-0.05, -0.02) | 0.467 |
| DBP, mmHg | -0.00 (-0.04, 0.05) | 0.872 | 0.02, (-0.04, 0.08) | 0.433 |
| MAP, mmHg | -0.01 (-0.05, 0.03) | 0.657 | -0.01 (-0.05, 0.07) | 0.709 |
| Hyperlipidemia | -0.58 (-1.58, 0.42) | 0.258 | -0.71 (-2.12, 0.70) | 0.322 |
| TG, mmol/L | -0.18 (-0.48, 0.13) | 0.265 | -0.15 (-0.59, 0.28) | 0.482 |
| LDL-C, mmol/L | -0.35 (-0.89, 0.19) | 0.204 | -0.43 (-1.188, 0.33) | 0.263 |
| HDL-C, mmol/L | -0.17 (-1.06, 1.40) | 0.785 | 0.06 (-1.67, 1.78) | 0.949 |
| TC, mmol/L | -0.33 (-0.81, 0.16) | 0.185 | -0.40 (-1.08, 0.28) | 0.249 |
| C-reactive protein, mg/dL | -0.06 (-0.13, -0.01) | 0.099 | -0.08 (-0.18, 0.02) | 0.121 |
| Microalbuminuria, mg/L | -0.01 (-0.03, 0.02) | 0.609 | -0.00 (-0.04, 0.04) | 0.964 |
| eGFR, mL/min/1.73 m<sup>2</sup> | 0.13 (0.08, 0.17) | **<0.001** | 0.15 (0.09, 0.22) | **<0.001** |
| IOP, mmHg | 0.23 (0.02, 0.43) | **0.029** | 0.32 (0.03, 0.60) | **0.030** |
| Axial length, mm | 0.20 (-0.27, 0.67) | 0.393 | 1.09 (0.43, 1.74) | **0.001** |

Abbreviations: GC-IPL, ganglion cell-inner plexiform layer; GCC, ganglion cell complex; 95%CI, 95% confidential interval; SD, standard deviation; SBP, systolic blood pressure; DBP, diastolic pressure; MAP, mean artery pressure; TG, triglyceride; LDL-C, low density lipoprotein cholesterol; HDL-C, high density lipoprotein cholesterol; TC, total cholesterol; IQR, interquartile range; eGFR, estimated glomerular filtration rate; IOP, intraocular pressure.

Table 4 shows the results of the multivariate linear regression. To reduce the effect of the presence of DR, participants with and without DR were analyzed separately. Model 1 was adjusted for the factors known to be relevant to macular GC–IPL and GCC thickness. These included age, sex, and axial length. Model 2 further adjusted for the parameters associated with macular GC–IPL and GCC thickness in the univariate model. In the patients without DR, the linear correlation between the eGFR and GC–IPL and GCC persisted in all the models (all, *p* < 0.01). In model 2, eGFR was linearly correlated with the macular GC-IPL (β = 0.07 [95% CI 0.02–0.012], *p* < 0.01) and GCC thickness (β = 0.09 [95% CI 0.03–0.016], *p* < 0.01). However, in the DR patients, there was no significant linear correlation between eGFR and macular GC-IPL or GCC thickness (all, *p* ≥ 0.05).

**Table 4.** Multivariate Linear Regression Analysis of the Relationship between Estimated Glomerular Filtration Rate (eGFR) and Thickness of Macular Ganglion Cell-inner Plexiform Layer or Ganglion Cell Complex.

| Parameters | No DR (n=1094) |  | DR (n=215) |  |
| --- | --- | --- | --- | --- |
|  | Coefficient (95%CI) | P-value | Coefficient (95%CI) | P-value |
| Macular GC-IPL thickness | 0.14 (0.09, 0.19) | **<0.001** | 0.10 (-0.05, 0.26) | 0.184 |
| Model 1<sup>†</sup> | 0.07 (0.02, 0.12) | **0.006** | 0.05 (-0.12, 0.23) | 0.551 |
| Model 2<sup>‡</sup> | 0.07 (0.02, 0.12) | **0.009** | 0.02 (-0.16, 0.20) | 0.831 |
| Macular GCC thickness | 0.18 (0.11, 0.24) | **<0.001** | 0.08 (-0.15, 0.31) | 0.489 |
| Model 1<sup>†</sup> | 0.10 (0.03, 0.16) | **0.004** | 0.00 (0.25, 0.25) | 0.990 |
| Model 2<sup>‡</sup> | 0.09 (0.03, 0.16) | **0.006** | -0.04 (-0.30, 0.22) | 0.777 |

Abbreviations: DR, diabetic retinopathy; 95%CI, 95% confidential interval; GC-IPL, ganglion cell-inner plexiform layer; GCC, ganglion cell complex.
<sup>†</sup>Adjusted for age, gender, axial length.
<sup>‡</sup>Further adjusted for hypertension (yes=1; no=0), intraocular pressure.

## 4. Conclusions

We have two main results: (1) The average macular GC–IPL and GCC thickness decreased in the CKD patients. (2) The eGFR was positively correlated with the average macular GC–IPL and GCC thickness independent of age, sex, axial length, and other potential confounding factors in patients without DR. Our results show a close association between retinal neurodegeneration and renal dysfunction and suggests that neurodegeneration progressed as renal function deteriorated. Thus, the potential for macular GC–IPL and GCC thickness to be indicative of early hyperglycemia-induced impairment in the eye and kidney in T2DM patients was supported.

We also have a few negative results. First, the macular GC–IPL and GCC thinning in CKD patients was found in every grid except the central grid. This could have been caused by the lower density of the ganglion cells in the fovea pit.^17^ Second, no significant linear correlation was found between macular GC–IPL and GCC thickness and the eGFR in the DR patients. One possible reason is the relatively small sample size (215) of DR patients. Another is that the increase in retinal thickness in DR patients might have obscured the GC–IPL and GCC thinning caused by the loss of ganglion cells. In the progression of DR, retinal capillary leakage would lead to retinal edema.^18^ Moreover, it has been hypothesized that ganglion cell degeneration could play a role in the blood-retinal barrier breakdown which could induce retinal edema in the development of DR.^19^ The results of the linear regressions indicate that the macular GC–IPL and GCC in the DR patients tended to be thicker than patients without DR. Before diabetic macular edema becomes clinically significant, morphologic changes might not be observable in OCT images. It is assumed that the evaluation of neurodegeneration through OCT might be more valuable for patients without DR.

The retinal and renal glomerular microvasculature have many similarities, and diseases of these organs have a common pathogenesis. However, few studies have evaluated the association between neurodegeneration and renal function in diabetic patients. Srivastav et al. reported that an increase in serum creatinine levels was associated with RNFL thinning in diabetic patients.^10^ However, the study included only 60 diabetic patients and considered few relative factors. Moreover, serum creatinine has limited value as a single indicator of renal function because more than half of the patients with an eGFR of <60 ml/min/1.73 m^2^ might have normal serum creatinine levels.^14^ The present study enrolled a large sample of T2DM patients in local communities and adjusted for multiple potential confounding factors. Thus, renal function was confirmed to be independently correlated with retinal neurodegeneration in diabetic patients.

There are also limitations. First, as was previously mentioned, the use of macular GC–IPL and GCC thickness to evaluate ganglion cell loss might have been confounded by the presence DR. Because of the uneven distribution of DR patients, further studies are necessary to investigate retinal neurodegeneration and its association with renal function in patients with various severity levels of DR. Second, we only analyzed the baseline data; thus, it is also necessary to further investigate the synchronization of retinal neurodegeneration and renal function decline. Follow-up visits are ongoing. Third, this was not a population-based or hospital-based study. The participants were recruited from communities voluntarily. A majority did not have DR or CKD; thus, the generalization of the findings to other diabetic patients should be considered with caution. Most patients with diabetes in China are located in communities, thereby this study still can represent a large population.

In summary, we found that retinal neurodegeneration progress with deterioration of renal function in Chinese T2DM patients. OCT is a non-invasive tool which can quantitatively evaluate GC-IPL and GCC thicknesses, but the evaluation is more valuable for patients without DR.

## Data Availability

The availability of all data referred to in the manuscript and note links below.

## Author contribution

W Huang and W Wang conceived and designed the study. X Gong, W Li, W Wei, L Wang, J Meng, Y Li, K Xiong and X Han collected and interpreted the data. X Gong and L Jin carried out the statistical analysis. X Gong wrote the manuscript. W Huang, W Wang, and W Li reviewed and edited the manuscript. All authors have seen the final version of the manuscript and approved it for publication.

## Acknowledgements and Funding

This work was supported by the National Natural Science Foundation of China (grand numbers 81570843; 81530028; 81721003), the Guangdong Province Science & Technology Plan (grant number 2014B020228002).

## Financial Disclosure

No potential conflicts of interest relevant to this article were reported.

## Notes

**Financial Conflict:** None

### Competing Interest Statement

The authors have declared no competing interest.

### Funding Statement

This study was funded by the Guangdong Province Science & Technology Plan (2014B020228002).

### Author Declarations

The Institute Ethics Committee of ZOC (2017KYPJ094)

## References

1. Drawz P RM. Chronic Kidney Disease. Ann Intern Med. 2015;162:C1–16.

2. Anders HJ Htib. CKD in Diabetes: Diabetic Kidney Disease Versus Nondiabetic Kidney Disease. Nat Rev Nephrol. 2018;14:361–77.

3. Zheng Y, Ley SH, Hu FB. Global Aetiology and Epidemiology of Type 2 Diabetes Mellitus and Its Complications. Nat Rev Endocrinol. 2018;14:88–98.

4. Saaddine JB, Honeycutt AA, Narayan KMV, et al. Projection of Diabetic Retinopathy and Other Major Eye Diseases Among People With Diabetes Mellitus: United States, 2005-2050. Archives of Ophthalmology 2008;126:1740–7.

5. Mohamed Q GMWT. Management of Diabetic Retinopathy: A Systematic Review. JAMA. 2007;298:902–16.

6. Barber AJ. Diabetic Retinopathy: Recent Advances towards Understanding Neurodegeneration and Vision Loss. Science China Life Sciences 2015;58:541–9.

7. Carpineto P TLAR. Neuroretinal Alterations in the Early Stages of Diabetic Retinopathy in Patients with Type 2 Diabetes Mellitus. Eye. 2016;30:673–9.

8. Pierro L, Iuliano L, Cicinelli MV, et al. Retinal Neurovascular Changes Appear Earlier in Type 2 Diabetic Patients. Eur J Ophthalmol. 2017;27:346–51.

9. Wong CW, Wong TY, Cheng C, Sabanayagam C. Kidney and Eye Diseases: Common Risk Factors, Etiological Mechanisms, and Pathways. Kidney Int. 2014;85:1290–302.

10. Srivastav K, Saxena S, Mahdi AA, et al. Increased Serum Urea and Creatinine Levels Correlate with Decreased Retinal Nerve Fibre Layer Thickness in Diabetic Retinopathy. Biomarkers 2015;20:470–3.

11. Li DY YWYY. What Is the Best Glomerular Filtration Marker to Identify People with Chronic Kidney Disease Most Likely to Have Poor Outcomes. Kidney Int. 2019;95:636–46.

12. Consultation WE. Appropriate Body-mass Index for Asian Populations and Its Implications for Policy and Intervention Strategies. The Lancet 2004;9403:157–63.

13. Li D, Yin W, Yi Y, et al. Development and Validation of a More Accurate Estimating Equation for Glomerular Filtration Rate in a Chinese population. Kidney Int. 2019;95:636–46.

14. Thomas MC BMSK. Diabetic Kidney Disease. Nat Rev Dis Primers. 2015;1.

15. Harding S, Greenwood R, Aldington S, et al. Grading and disease management in national screening for diabetic retinopathy in England and Wales. Diabet Med. 2003;20:965–71.

16. Knight ORJ, Girkin CA, Budenz DL, et al. Effect of Race, Age, and Axial Length on Optic Nerve Head Parameters and Retinal Nerve Fiber Layer Thickness Measured by Cirrus HD-OCT. Archives of Ophthalmology 2012;130:312–8.

17. Chui TY SHBS. Adaptive-optics Imaging of Human Cone Photoreceptor Distribution. J Opt Soc Am A Opt Image Sci Vis. 2008;25:3021–9.

18. Cheung N MPWT. Diabetic Retinopathy. Lancet. 2010;9735:124–36.

19. Fdaj CJ. Early Breakdown of the Blood-retinal Barrier in Diabetes. Br J Ophthalmol. 1975;59:649–56.

